# Physical Frailty Accelerates Multimorbidity Accumulation Across Distinct Disease Patterns

**DOI:** 10.64898/2026.10.15.26365629

**Authors:** Yaqi Huang, Zixin Hu, Meng Hao, Xiangnan Li, Yulong Tang, Shuai Jiang, Xiaofeng Wang, Li Han, Jiucun Wang, Yi Li, Hui Zhang

**Affiliations:** Human Phenome Institute, School of Life Sciences, Huadong Hospital, Fudan University, Shanghai, 200433, China; Artificial Intelligence Innovation and Incubation Institute, Fudan University, Shanghai 201111, China; Huadong Hospital, Shanghai Key Laboratory of Clinical Geriatric Medicine, Shanghai Medical College, Fudan University, Shanghai, 200040, China; Section of Epidemiology and Population Sciences, Department of Medicine, Baylor College of Medicine, Houston, Texas, 77030, USA; Department of Biomedical Genetics, University of Rochester, Rochester, New York, 14627, USA; Department of Vascular Surgery, Shanghai Key Laboratory of Vascular Lesion Regulation and Remodeling, Shanghai Pudong Hospital, Fudan University Pudong Medical Center, Shanghai, 201399, China; School of Global Health, Chinese Center for Tropical Diseases Research, Shanghai Jiao Tong University School of Medicine, Shanghai, 200025, China

**Keywords:** Frailty, Multimorbidity, Multimorbidity accumulation, Disease patterns, Cohort study

## Abstract

**Background:** Emerging evidence has linked frailty to individual chronic diseases, but whether it predicts a broad disease spectrum and accelerates multimorbidity accumulation over time remains unknown. This study aimed to systematically evaluate the associations of physical frailty with the burden of incident disease across a wide range of chronic conditions, and with the long-term rates of multimorbidity accumulation.

**Methods:** In this prospective cohort study of 466,864 UK Biobank participants (baseline mean age 56.51 ± 8.08 years), frailty status was assessed using the Fried frailty phenotype and categorized as robust, pre-frailty, or frailty. Chronic diseases were defined based on International Classification of Diseases (ICD)-10 codes, and disease patterns were identified using network analysis. Cox proportional hazards models were used to examine the associations of frailty status with incident chronic diseases, and linear mixed models were used to assess the annual rate of chronic disease accumulation.

**Results:** During a median follow-up of 13.8 years (IQR: 13.1-14.5), 536 incident chronic diseases across multiple systems were recorded. Physical frailty was associated with a significantly increased risk of 458 incident diseases (Bonferroni-corrected P < 0.05/536). In the overall cohort, compared with robust participants, those with pre-frailty (β = 0.266, 95% confidence interval [CI]: 0.264-0.267) and frailty (β = 0.941, 95% CI: 0.936-0.946) exhibited significantly faster rates of multimorbidity accumulation. This accelerated multimorbidity accumulation was consistently observed across strata of baseline disease burden, including participant with 0 (β = 0.644, 95% CI: 0.635-0.653), 1 (β =0.748, 95% CI: 0.727-0.768), ≥ 2 (β =0.940, 95% CI: 0.933-0.947) chronic conditions at baseline. Similar associations were also found across distinct disease patterns, including neurodegenerative, metabolic, and psychiatric patterns.

**Conclusions:** Physical frailty predicts a broad range of incident diseases and is associated with accelerated multimorbidity accumulation. Incorporating frailty assessment into prevention strategies may help mitigate the growing burden of multimorbidity.

## Introduction

Population ageing poses a major challenge to health and social care systems worldwide[1]. Ageing increases vulnerability to the development of multiple chronic diseases and is regarded as the primary risk factor for multimorbidity[2, 3]. Multimorbidity, defined as the presence of at least two chronic conditions, affects approximately 37.2% of adults worldwide, and is associated with increased risks of multiple adverse health outcomes[4–6]. Moreover, multimorbidity imposes a large economic burden on health systems and society, with annual healthcare costs of multimorbidity ranging from 800 to 150,000 dollars per person depending on disease combinations and countries[7]. Therefore, given its high prevalence and substantial burden, multimorbidity has become a growing public health challenge globally, highlighting the need to identify factors that influence chronic disease accumulation[4, 8, 9].

Physical frailty is a clinical syndrome characterized by reduced physiologic reserve across multiple organ systems, and increased susceptibility to stressors[10]. It is associated with increased risks of numerous adverse health outcomes, including falls, hospitalization, disability, and mortality[11]. Critically, frailty is potentially modifiable through targeted interventions, such as physical activity and nutritional support[12–14]. Thus, integrating frailty assessment into clinical practice may facilitate earlier identification of high-risk individuals, and support a shift from disease treatment toward prevention[15]. Despite the well-established association between frailty and multimorbidity, longitudinal evidence on their relationship has several critical gaps[16–18]. For example, previous studies have predominantly focused on a limited range of chronic conditions when examining the association with frailty. A systematic review reported that only 4 to 28 different conditions were considered in assessing multimorbidity, suggesting the restricted disease coverage in existing literature[16]. Additionally, most studies have generally operationalized multimorbidity as a binary status (presence or absence), or as simple disease counts, and thus may provide limited information on the process of disease accumulation over time[19]. This distinction is clinically relevant because the rate of accumulation may better capture dynamic disease progression, and enable earlier identification of individuals with accelerated health deterioration[20, 21]. Moreover, whether frailty differentially influences the development of distinct multimorbidity patterns (clusters of diseases that tend to co-occur) across organ systems remains poorly understood[16, 22, 23].

To address these gaps, we conducted a prospective cohort study using data from UK Biobank. The main aim of this study was to systematically examine whether physical frailty predicts a broad spectrum of incident chronic diseases and the longitudinal rate of multimorbidity accumulation across multiple systems. By providing a comprehensive, system-level assessment of disease burden over time, this study may help identify individuals at high risk of accelerated multimorbidity progression and guide targeted strategies for earlier prevention and management.

## Methods

### Study Population

The UK Biobank is a large prospective population-based cohort comprising more than 500,000 individuals from England, Scotland and Wales. During the baseline assessment between 2006 and 2010, data on lifestyle factors, environmental exposures, medical history, physical measurements and biological samples were collected[24]. Participants with missing data required for physical frailty assessment were excluded. Finally, a total of 466,864 participants were included.

### Assessment of Physical Frailty

Physical frailty was assessed using the five-component frailty phenotype proposed by Fried et al[25]. The phenotype comprises five criteria: unintentional weight loss, exhaustion, low physical activity, slow gait speed and weakness. In detail, unintentional weight loss was assessed by the question “Compared with one year ago, has your weight changed?” Unintentional weight loss was defined as reporting “yes, lost weight”. Exhaustion was assessed by the question “Over the past two weeks, how often have you felt tired or had little energy?” Participants were considered to meet the exhaustion criterion if they responded “more than half the days” or “nearly every day”. Physical activity was assessed by the question “In the last 4 weeks did you spend any time doing light do-it-yourself (DIY) activity, heavy DIY activity, or strenuous sports?” and “How many times in the last 4 weeks did you do light DIY?” Participants were considered to meet the low physical activity criterion if they responded “none of the above” or “light activity with a frequency of once per week or less”. Gait speed was assessed by the question “How would you describe your usual walking pace?” Participants were considered to meet the slow gait speed criterion if they responded “slow pace.” Weakness was assessed using the maximum grip strength recorded for either hand. Participants were considered to meet the weakness criterion if their grip strength was below the sex- and body mass index-adjusted cutoffs proposed by Fried[25]. Participants were categorized as “frailty” if they met at least three of the five criteria, “pre-frailty” if they met one or two criteria, and “robust” if they met none.

### Definition of Chronic Diseases and Multimorbidity

In the UK Biobank, disease diagnoses were obtained by linkage to UK electronic health records, and defined using the International Classification of Diseases (ICD)-10 codes. Incident diseases were defined as diagnoses occurring after the date of participants’ baseline assessment. Moreover, the follow-up was censored at the earliest occurrence of disease diagnosis, death, loss to follow-up, or end of follow-up (October 31, 2022) for each chronic disease. Follow-up observations were discontinued at death, loss to follow-up, or the administrative censoring date.

Following previous method proposed by Deng et al.[26], disease endpoints were constructed through the FinnGen disease endpoints framework (https://www.finngen.fi/en/researchers/clinical-endpoints) and adhered to FinnGen’s quality control. Specifically, we incorporated pre-defined conditions based on exclusion criteria for the particular diseases. For each chronic disease, participants with prevalent diagnosis records were excluded separately. Disease endpoints were further excluded if they met any of the following criteria: (1) fewer than 400 incident cases during the follow-up (approximately 0.1% of the total population)[27]; (2) age- or sex-specific diseases; (3) infectious diseases; (4) benign tumors (while malignant neoplasms were retained). As a result, a total of 536 chronic diseases were included in the analysis. Detailed information of included chronic diseases is provided in **Supplementary Table 1**. Multimorbidity was further defined as the cumulative number of chronic diseases.

### Multimorbidity patterns

Network analysis was performed to identify non-random co-occurrence patterns among chronic conditions. We constructed a weighted, undirected disease network where each node represented a chronic disease and each edge indicated the association strength between two chronic conditions. Partial correlation was used to quantify these associations because it additionally controlled the effects of all other conditions in the network[28]. Larger partial correlation coefficients indicated stronger associations. Edges with low partial correlation coefficients below 0.01 were excluded from subsequent clustering. To investigate the community structure within the multimorbidity network, the Louvain algorithm was applied with a resolution parameter of 0.9 based on suitable modularity size and cluster interpretability[23]. The Louvain algorithm was computationally efficient to detect communities in large-scale networks. Communities identified by the algorithm were interpreted as disease clusters with stronger within-cluster associations.

### Covariates

The covariates comprised sociodemographic characteristics and lifestyle factors. The sociodemographic characteristics included age, gender, race/ethnicity, education and Townsend deprivation index (TDI). Race/ethnicity was categorized as White and non-White. In the UK Biobank, participants reporting British, Irish, or any other White background were classified as White, whereas all other backgrounds were classified as non-White. Education was categorized as less than high school, high school or equivalent, and college or above. Participants reporting a college or university degree were classified as having college or above education; those reporting none of the listed qualifications were classified as having less than high school education; and all other qualifications were classified as high school or equivalent. Lifestyle factors included body mass index (BMI), alcohol consumption (ever/never), smoking status (ever/never) and insomnia symptoms (never, sometimes, usually).

### Statistical Analyses

Baseline characteristics were summarized using mean and standard deviation (SD) for continuous variables, and frequency (%) for categorical variables, respectively. Differences across groups were analyzed by chi-square test or ANOVA based on the data type. Participants were categorized into three groups according to baseline chronic disease burden as disease-free (0 disease), single chronic condition (1 disease), and multimorbidity (≥ 2 diseases).

Cox proportional hazards regression models were used separately to examine the relationships between physical frailty and a broad disease spectrum, with adjustment for age, gender, race/ethnicity, education, TDI, BMI, alcohol consumption, smoking status, and insomnia symptom. Statistical significance was determined after Bonferroni correction for multiple testing (P < 0.05/536). Linear mixed models with participant-specific random intercepts were applied to assess the association between baseline physical frailty and the rate of chronic disease accumulation. Interactions between frailty and follow-up time were included as fixed effects, which estimated the differences in the average annual increase in the number of chronic diseases. Follow-up time was represented as completed years since baseline assessment (1, 2, … years) in the linear mixed models. Robust participants served as the reference group. Model 1 was the crude model; Model 2 was adjusted for age, gender, race/ethnicity, education, and TDI; and Model 3 was additionally adjusted for BMI, alcohol consumption, smoking status, and insomnia symptom. Moreover, when analyzing the accumulation of specific disease systems, the number of chronic diseases outside that system was additionally included in the model. All analyses were repeated across subgroups with different baseline disease burdens (disease-free, single chronic condition, and multimorbidity). P values were 2-sided and less than 0.05 were considered statistically significant.

We further examined effect modification by age and sex by including three-way interactions among physical frailty status, follow-up time, and age group (< 50, 50-64, and ≥ 65 years), or sex (female and male). Sensitivity analyses were performed to assess the robustness of the main findings: (1) excluding participants who developed any incident disease within 3 years after baseline; and (2) restricting analyses to participants with at least 5 years of follow-up. All analyses were performed using R statistical software, version 4.4.1.

## Results

### Characteristics of Study Population

**Table 1** presents the baseline characteristics of the study population according to baseline chronic disease burden. The analysis included 466,864 participants with complete data for all five physical frailty indicators. Of these, the mean baseline age was 56.51 (8.08) years, and 253,687 (54.34%) were female. 273,498 individuals (58.58%) were classified as robust, 176,403 (37.78%) as pre-frailty, and 16,963 (3.63%) as frailty. According to baseline chronic disease burden, 236,403 individuals (50.64%) were free of chronic disease, 35,676 (7.64%) had a single chronic condition, and 194,785 (41.72%) had baseline multimorbidity.

**Table 1.** Characteristics of study population at baseline.

| Characteristics | Total | No. of chronic diseases at baseline |  |  | P-value |
| --- | --- | --- | --- | --- | --- |
|  |  | Disease-free | Single disease | Multimorbidity |  |
| Sample, N (%) | 466864 (100.00) | 236403 (50.64) | 35676 (7.64) | 194785 (41.72) |  |
| Age, years, M (SD) | 56.51 (8.08) | 55.30 (8.10) | 56.26 (8.00) | 58.02 (7.81) | 0.00E+00 |
| Age Groups, N (%) |  |  |  |  | 0.00E+00 |
| < 50 years | 109592 (23.47) | 66559 (28.15) | 8589 (24.08) | 34444 (17.68) |  |
| 50-64 years | 269026 (57.62) | 135019 (57.11) | 20831 (58.39) | 113176 (58.10) |  |
| ≥ 65 years | 88246 (18.90) | 34825 (14.73) | 6256 (17.54) | 47165 (24.21) |  |
| Gender, N (%) |  |  |  |  | 0.00E+00 |
| Female | 253687 (54.34) | 125251 (52.98) | 24582 (68.90) | 103854 (53.32) |  |
| Male | 213177 (45.66) | 111152 (47.02) | 11094 (31.10) | 90931 (46.68) |  |
| Race/Ethnicity, N (%) |  |  |  |  | 8.54E-06 |
| White | 442782 (94.84) | 223888 (94.71) | 33974 (95.23) | 184920 (94.94) |  |
| Others | 24082 (5.16) | 12515 (5.29) | 1702 (4.77) | 9865 (5.06) |  |
| Education, N (%) |  |  |  |  | 0.00E+00 |
| < High school | 76444 (16.37) | 29182 (12.34) | 5682 (15.93) | 41580 (21.35) |  |
| High school or equivalent | 232179 (49.73) | 116248 (49.17) | 18274 (51.22) | 97657 (50.14) |  |
| ≥ College or above | 154126 (33.01) | 89225 (37.74) | 11432 (32.04) | 53469 (27.45) |  |
| TDI, Median (IQR) | -2.19 (-3.67, 0.42) | -2.34 (-3.76, 0.12) | -2.23 (-3.67, 0.32) | -1.99 (-3.55, 0.82) | 0.00E+00 |
| BMI, kg/m2, M (SD) | 27.39 (4.77) | 26.89 (4.48) | 27.01 (4.64) | 28.07 (5.04) | 0.00E+00 |
| Alcohol consumption, N (%) |  |  |  |  | 0.00E+00 |
| Ever | 430961 (92.31) | 221799 (93.82) | 33129 (92.86) | 176033 (90.37) |  |
| Never | 35535 (7.61) | 14476 (6.12) | 2525 (7.08) | 18534 (9.52) |  |
| Smoking status, N (%) |  |  |  |  | 0.00E+00 |
| Ever | 210554 (45.10) | 99437 (42.06) | 15621 (43.79) | 95496 (49.03) |  |
| Never | 254861 (54.59) | 136367 (57.68) | 19958 (55.94) | 98536 (50.59) |  |
| Insomnia |  |  |  |  | 0.00E+00 |
| Never | 113634 (24.34) | 64793 (27.41) | 7919 (22.20) | 40922 (21.01) |  |
| Sometimes | 221667 (47.48) | 114590 (48.47) | 17460 (48.94) | 89617 (46.01) |  |
| Usually | 131296 (28.12) | 56891 (24.07) | 10275 (28.80) | 64130 (32.92) |  |
| Frailty status |  |  |  |  | 0.00E+00 |
| Robust | 273498 (58.58) | 152527 (64.52) | 21371 (59.90) | 99600 (51.13) |  |
| Pre-frailty | 176403 (37.78) | 79940 (33.82) | 13396 (37.55) | 83067 (42.65) |  |
| Physical frailty | 16963 (3.63) | 3936 (1.66) | 909 (2.55) | 12118 (6.22) |  |
| Incident chronic diseases, Number, M (SD) | 13.45 (13.73) | 8.50 (10.67) | 11.93 (12.13) | 19.73 (14.74) | 0.00E+00 |

Missing values were as follows: education 4,115 (0.88%); TDI 678 (0.15%); alcohol consumption 368 (0.08%); smoking status 1,449 (0.31%); and insomnia, 267 (0.06%). TDI: Townsend Deprivation Index; BMI: body mass index; M: mean; SD: standard deviation.

### Frailty and Risk of Incident Chronic Diseases Across a Broad Disease Spectrum

We examined the associations between physical frailty and the risks of 536 incident chronic diseases. A total of 435 diseases (81.16%) and 458 (85.45%) diseases were significantly associated with pre-frailty and frailty, respectively, after Bonferroni correction (P < 0.05/536 = 9.33 × 10^-5^; **Figure 1**). These associated diseases spanned multiple organ systems, demonstrating the broad and multisystem associations of frailty (**Supplementary Table 2**).

**Table 2.** Association between physical frailty and the annual rate of total chronic disease accumulation.

|  |  | **Model 1** |  | **Model 2** |  | **Model 3** |  |
| --- | --- | --- | --- | --- | --- | --- | --- |
|  |  | **β (95% CI)** | **P-value** | **β (95% CI)** | **P-value** | **β (95% CI)** | **P-value** |
| Total | Robust | Ref |  | Ref |  | Ref |  |
|  | Pre-frailty | 0.266 (0.264-0.267) | 0.00E+00 | 0.266 (0.264-0.268) | 0.00E+00 | 0.266 (0.264-0.267) | 0.00E+00 |
|  | Physical frailty | 0.941 (0.936-0.946) | 0.00E+00 | 0.941 (0.936-0.946) | 0.00E+00 | 0.941 (0.936-0.946) | 0.00E+00 |
| Baseline disease free | Robust | Ref |  | Ref |  | Ref |  |
|  | Pre-frailty | 0.178 (0.176-0.180) | 0.00E+00 | 0.178 (0.176-0.181) | 0.00E+00 | 0.178 (0.176-0.181) | 0.00E+00 |
|  | Physical frailty | 0.644 (0.635-0.653) | 0.00E+00 | 0.644 (0.635-0.653) | 0.00E+00 | 0.644 (0.635-0.653) | 0.00E+00 |
| Baseline single chronic disease | Robust | Ref |  | Ref |  | Ref |  |
|  | Pre-frailty | 0.226 (0.219-0.232) | 0.00E+00 | 0.226 (0.220-0.233) | 0.00E+00 | 0.226 (0.220-0.233) | 0.00E+00 |
|  | Physical frailty | 0.748 (0.727-0.768) | 0.00E+00 | 0.748 (0.728-0.769) | 0.00E+00 | 0.748 (0.727-0.768) | 0.00E+00 |
| Baseline multimorbidity | Robust | Ref |  | Ref |  | Ref |  |
|  | Pre-frailty | 0.304 (0.300-0.307) | 0.00E+00 | 0.304 (0.301-0.307) | 0.00E+00 | 0.304 (0.301-0.307) | 0.00E+00 |
|  | Physical frailty | 0.940 (0.934-0.947) | 0.00E+00 | 0.941 (0.934-0.947) | 0.00E+00 | 0.940 (0.933-0.947) | 0.00E+00 |

Model 1 was crude model; Model 2 was adjusted for age, gender, race/ethnicity, education, and TDI; Model 3 were additionally adjusted for BMI, alcohol consumption, smoking status, and insomnia symptom. TDI: Townsend Deprivation Index; BMI: body mass index; CI: confidence interval.

**Figure 1.** Associations between physical frailty and a broad spectrum of incident chronic diseases. Associations between physical frailty and 536 incident diseases were estimated by the Cox proportional hazards regression models adjusted for age, sex (female or male), race/ethnicity (White or others), education (less than high school, high school or equivalent, or college or above), TDI, BMI, alcohol consumption (yes or no), smoking status (ever or never), and insomnia symptom (never, sometimes or usually). Lines represent hazard ratios (HRs) for chronic diseases associated with pre-frailty or frailty. Diseases are grouped according to network analysis, with different colors representing different clusters. Within each disease cluster, diseases are ordered according to the magnitude of HRs. Gray lines indicate associations that did not reach statistical significance after Bonferroni correction (P < 0.05/536 = 9.3 × 10 □ □). Triangles indicate HRs greater than 6 for frailty and 2 for pre-frailty that were truncated for visualization; the complete HRs and 95% CIs are provided in Supplementary Table 2. Selected diseases of clinical importance or novelty are annotated. COPD: chronic obstructive pulmonary disease. T2D: Type 2 diabetes. TDI: Townsend Deprivation Index. BMI: body mass index.

Among these associations, postviral fatigue syndrome showed the highest HRs for both pre-frailty and frailty, with HRs of 3.294 (95% CI: 2.844-3.815) and 14.030 (95% CI: 11.480-17.145), respectively. This was followed by fibromyalgia, with corresponding HRs of 2.693 (95% CI: 2.480-2.925) and 10.147 (95% CI: 9.120-11.291), respectively (**Figure 1**). Additionally, Our findings also confirmed previously reported associations between frailty and several chronic conditions, such as Parkinson’s disease (HR = 2.542, 95% CI: 2.206-2.929), heart failure (HR = 2.840, 95% CI: 2.681-3.008), irritable bowel syndrome (HR = 2.839, 95% CI: 2.622-3.074), and abdominal aortic aneurysm (HR = 2.365, 95% CI: 2.000-2.796). Notably, our results also revealed many novel associations. For example, frailty was significantly associated with the risk of bronchiectasis (HR = 4.423, 95% CI: 4.009-4.880), with similar associations observed for idiopathic pulmonary fibrosis (HR = 3.105, 95% CI: 2.721-3.544), dialysis (HR = 5.711, 95% CI: 4.591-7.104), peripheral artery disease (HR = 3.972, 95% CI: 3.635-4.339), and cardiomyopathy (HR = 2.419, 95% CI: 2.079-2.816). We also examined these associations across subgroups with different baseline disease burdens (**Supplementary Tables 3-5**). A total of 246, 140, and 440 chronic diseases showed significant associations with frailty after Bonferroni correction among participants in baseline disease-free status, single chronic condition, and multimorbidity, respectively.

### Frailty and Rate of Total Chronic Disease Accumulation

The coefficients for differences in annual rates of chronic disease accumulation according to frailty status are shown in **Table 2**. After full adjustment, both pre-frailty and frailty were associated with faster total chronic disease accumulation in all participants, with additional annual increases of 0.266 (95% CI: 0.264-0.267) and 0.941 (95% CI: 0.936-0.946) chronic diseases per year, respectively, compared with robust participants. The corresponding associations remained evident across baseline disease burden groups. Among baseline disease-free participants, pre-frailty and frailty were associated with 0.178 (95% CI: 0.176-0.181) and 0.644 (95% CI: 0.635-0.653) additional annual chronic diseases, respectively. The corresponding β coefficients were 0.226 (95% CI: 0.220-0.233) and 0.748 (95% CI: 0.727-0.768) among participants with single chronic condition at baseline, and 0.304 (95% CI: 0.301-0.307) and 0.940 (95% CI: 0.933-0.947) among those with baseline multimorbidity.

**Figure 2** illustrates the longitudinal trajectories of chronic disease accumulation over follow-up according to frailty status across baseline disease-burden groups. Among all participants, those with pre-frailty and frailty had 3.973 (95% CI: 3.927-4.019) and 16.015 (95% CI: 15.892-16.137) more accumulated chronic diseases than robust participants during 15 years of follow-up, respectively. Among baseline disease-free participants, the corresponding differences were 2.183 (95% CI: 2.138-2.229) and 8.346 (95% CI: 8.178-8.515), respectively. Similar patterns were observed among participants with a single chronic condition and multimorbidity at baseline, with larger absolute differences in accumulated disease numbers. The longitudinal trajectories of chronic disease accumulation in Model 1 and Model 2 are shown in **Supplementary Figures 1-2**.

**Figure 2.** Association between physical frailty and the annual rate of total chronic disease accumulation according to baseline disease burden. Linear mixed models were adjusted for age, sex (female or male), race/ethnicity (White or others), education (less than high school, high school or equivalent, or, college or above), TDI, BMI, alcohol consumption (yes or no), smoking status (ever or never), and insomnia symptom (never, sometimes or usually). Analysis was conducted across four groups: all participants, baseline disease-free, single baseline condition, and baseline multimorbidity. Data represent average predicted chronic disease counts with 95% CIs. TDI: Townsend Deprivation Index. BMI: body mass index. CIs: confidence intervals.

### Frailty and System-specific Patterns of Chronic Disease Accumulation

Network analysis initially identified 31 multimorbidity patterns, of which 15 major patterns with relatively large sizes (≥ 15 diseases) and greater clinical importance were retained in downstream analyses (**Supplementary Table 6**). Similar trends were observed for the accumulation of specific disease systems among all participants (**Figure 3, Supplementary Tables 7-9**). Both pre-frailty and frailty were associated with faster rates of disease accumulation, with larger coefficients generally observed in the frail individuals. For example, the β coefficients for pre-frailty and frailty were 0.011 (95% CI: 0.011-0.012) and 0.026 (95% CI: 0.026-0.027) for musculoskeletal joint diseases, 0.013 (95% CI: 0.013-0.013) and 0.044 (95% CI: 0.044-0.045) for psychiatric diseases, and 0.005 (95% CI: 0.004-0.005) and 0.017 (95% CI: 0.016-0.017) for metabolic diseases, respectively. Similar patterns between baseline frailty and accelerated system-specific disease accumulation were observed across different baseline disease burden groups.

**Figure 3.** Heatmap of association between physical frailty and the annual rate of chronic disease accumulation across multiple disease systems. The Louvain algorithm was applied to identify the community structure within the multimorbidity network. Linear mixed models were fully adjusted for age, sex (female or male), race/ethnicity (White or others), education (less than high school, high school or equivalent, or, college or above), TDI, BMI, alcohol consumption (yes or no), smoking status (ever or never), and insomnia symptom (never, sometimes or usually). P values were two sided (*P □ <□0.05, **P□<□0.01, and ***P□< □0.001). No adjustments were made for multiple comparisons.

### Stratified Analyses in Subgroups

Stratified analyses were conducted according to age group (< 50, 50-64, and ≥ 65 years) and sex (female and male). The associations between physical frailty status and chronic disease accumulation were generally consistent across strata, but the magnitude of the associations varied across subgroups (**Supplementary Tables 10-11**). In age-stratified analyses, the excess rate of total chronic disease accumulation was greater in older participants (P for interaction < 0.001). Similar patterns were observed for some specific disease systems, such as metabolic, respiratory, and renal-urinary diseases, while some specific systems showed different age-specific patterns. For instance, larger coefficients were observed in age group < 50 years for psychiatric, and gastrointestinal diseases. In sex-stratified analyses, the excess rate of total chronic disease accumulation was faster in females (P value for interaction < 0.001). However, the sex-specific patterns varied across disease systems, with stronger associations among women for some systems, such as musculoskeletal joint and gastrointestinal diseases, and stronger associations among men for others, such as cardiovascular and psychiatric diseases.

### Sensitivity Analyses

These associations remained consistent in sensitivity analyses that excluded participants with incident diseases during the first 3 years of follow-up, or restricted analyses to participants with at least 5 years of follow-up. The detailed results of total and system-specific chronic disease accumulation among all participants and across baseline disease burden groups are presented in **Supplementary Tables 12-13**.

### Discussion

In this prospective population-based study, physical frailty was associated with a broad range of incident chronic diseases and an accelerated rate of multimorbidity accumulation over long-term follow-up. These associations were evident across multiple disease systems and levels of baseline chronic disease burden, suggesting that frailty represents an early marker of multisystem multimorbidity accumulation. Together, these results support the integration of frailty assessment into strategies for multimorbidity prevention and healthy ageing in the general population.

Previous longitudinal studies have provided important evidence that frailty is associated with the subsequent presence of multimorbidity, disease counts, or specific disease patterns[17, 29]. However, these outcomes represent static descriptions of multimorbidity and may not capture the temporal progression of disease burden[30]. Moreover, different operational definitions of multimorbidity (including count-based and cluster-based approaches) can influence prevalence estimates and associations with health outcomes[31]. Our findings extend the existing literature on the relationship between physical frailty and multimorbidity. First, by systematically evaluating a broad spectrum of chronic diseases rather than limiting to prespecified outcomes, we confirmed previously reported associations (e.g., Parkinson’s disease[32]), and identified multiple novel associations (e.g., idiopathic pulmonary fibrosis). Second, by quantifying the rate of chronic disease accumulation, we shifted the analytical focus from binary status or disease patterns to the dynamic pace of accumulation over time. This approach not only identifies physical frailty as a predictor of future multimorbidity burden but also illustrates how it relates to the rate of multimorbidity development. Collectively, these findings provide population □ based evidence supporting the integration of frailty assessment into broader chronic disease prevention and management strategies, and may inform earlier preventive interventions for individuals at risk of accelerated multimorbidity progression.

Beyond the breadth of frailty □ associated diseases, our findings also provide insights into potential mechanisms underlying these multisystem associations. Associations between physical frailty and disease accumulation were observed across multiple disease systems, supporting the interpretation of frailty as a marker of multisystem vulnerability rather than a single disease domain[33]. The particularly pronounced accumulation of musculoskeletal diseases among frail individuals is consistent with the central role of musculoskeletal function in the Fried frailty phenotype[34], and may reflect shared biological pathways, including chronic low-grade inflammation, altered calcium and phosphate homeostasis and hormonal dysregulation[35]. We also found significant associations with psychiatric and neurological diseases, which are consistent with previous studies. These findings suggest that frailty is associated with atrophy in subcortical regions, which are closely involved in cognitive function and memory[36]. Potential mechanisms may include mitochondrial dysfunction, cellular apoptosis, altered cholesterol metabolism and changes in particular protein levels[37]. In addition, shared pathophysiological pathways between physical frailty and metabolic diseases may contribute to the impairments in glucose transport across cellular membranes and fatty acid transport across mitochondrial membranes[38, 39].

Interaction analyses revealed that the association of physical frailty with multimorbidity accumulation was modified by sex and age. Consistent with previous evidence that women suffer more multimorbidity than men and bear a larger burden of noncommunicable diseases[40], our study further suggests that women with frailty may also experience a faster rate of chronic disease accumulation. Interestingly, we observed that the accumulation rate of psychiatric and gastrointestinal diseases was faster among relatively younger frail individuals. This may partly reflect the fact that mental stress in earlier adulthood is more likely to be accompanied by elevated inflammatory levels and impaired immune function[41]. In addition, epidemiological evidence suggests that the prevalence of functional gastrointestinal disorders, such as dyspepsia and irritable bowel syndrome, tends to decrease with age[42]. These findings highlight that chronic disease progression may already occur in earlier stages of adulthood, demonstrating the importance of early identification and targeted prevention among younger individuals with frailty.

These findings have several implications. By systematically assessing associations across a broad spectrum of chronic diseases, our study provides a comprehensive atlas that may offer a systems-level framework for understanding the multisystem associations of frailty. Moreover, the observation of accelerated disease accumulation even among pre-frail individuals suggests that prevention strategies should not be limited to those with established frailty, but should extend to the pre-frailty stage. Both pre-frail and frail individuals may benefit from earlier multisystem assessment rather than disease-specific approaches. Finally, the consistent associations observed among participants who were disease-free at baseline suggest that physical frailty is not merely a concern in individuals with established multimorbidity, but may represent an earlier target in the general population. Collectively, these findings support a shift from single-disease care towards integrated multimorbidity prevention, and may inform strategies to promote healthy ageing.

Our study has several strengths. The analysis covered a broad range of diseases across multiple organ systems, providing a comprehensive assessment of multimorbidity development. We evaluated these associations not only in the overall population but also across different baseline chronic disease-burden groups. This design could assess the consistency of the findings and identify how pre-existing disease burden may influence frailty-related disease progression. Additionally, we examined the association between physical frailty and the rate of chronic disease accumulation, a dynamic measure proposed as a proxy for accelerated ageing[19]. Limitations should also be acknowledged. First, although we adjusted for a comprehensive set of confounders, residual confounding by unmeasured factors cannot be fully excluded. Second, frailty was assessed only at baseline, and potential changes in frailty status over time were not captured. Third, the number of diseases included in each organ system was not identical, and the effects across systems were not directly comparable. Fourth, the UK Biobank is predominantly of White European ancestry, which may limit the generalizability of our findings to other populations. Fifth, our analyses focused on the total number of incident diseases rather than the sequence or temporal order of disease onsets, which may provide additional insights into multimorbidity progression patterns. Finally, all chronic conditions were assigned equal weight without accounting for disease severity or stage, which may influence the rate of disease accumulation.

## Conclusion

In this large-scale, prospective cohort study, physical frailty was associated with a broad spectrum of incident chronic diseases and faster multimorbidity accumulation, with broadly consistent associations across multimorbidity patterns and baseline disease strata. Our findings suggest that frailty assessment could serve as an early-warning signal for accelerated health deterioration and enable earlier intervention. Integrating frailty assessment into primary care and community settings may help identify individuals at increased risk of accelerated chronic disease accumulation and inform risk-stratified prevention strategies. Future research should examine whether interventions aimed at preventing or reversing frailty can effectively reduce multimorbidity incidence and progression across diverse populations.

## Data Availability

The data used in this study were obtained from the UK Biobank (https://www.ukbiobank.ac.uk/). These data are available to qualified researchers upon application to the UK Biobank and are subject to its access procedures and restrictions.

## Acknowledgements

The data and samples used for this research were obtained from the UK Biobank. We would like to thank the workers, researchers, and participants involved in these cohorts. This research was conducted using the UK Biobank Resource under Application Number 103791.

## Data sharing statement

The dataset analyzed in the study was from the UK Biobank are available upon application at www.ukbiobank.ac.uk/register-apply. The datasets generated and analyzed during the current study are available from the corresponding author on reasonable request.

## Ethics approval and consent to participate

All participants provided electronic informed consent. The UK Biobank study received approval from the National Information Governance Board for Health and Social Care and the National Health Service North West Multi-Centre Research Ethics Committee.

## Authors’ contributions

Zhang H, Huang Y, Hu Z, and Li Y designed and conducted the research. Huang Y, Han L, Hao M, Li X, Tang Y, Hu Z, Jiang S, Wang X, Wang J, and Zhang H performed acquisition, analysis, or interpretation of data. Huang Y drafted the manuscript. Zhang H, and Li Y supervised the whole study. Zhang H, Huang Y, and Li Y had primary responsibility for the final content, and all authors read and approved the final manuscript.

## Legends

**Supplementary Table 1. Characteristics and definition of incident chronic diseases**.

**Supplementary Table 2. Associations between physical frailty and the risk of incident chronic diseases in the overall population**.

**Supplementary Table 3. Associations between physical frailty and the risk of incident chronic diseases among baseline disease-free participants**.

**Supplementary Table 4. Associations between physical frailty and the risk of incident chronic diseases among participants with baseline single chronic condition**.

**Supplementary Table 5. Associations between physical frailty and the risk of incident chronic diseases among participants with baseline multimorbidity**.

**Supplementary Table 6. Data-driven multimorbidity patterns identified by network analysis**.

**Supplementary Table 7. Associations between physical frailty and the annual rate of system-specific chronic disease accumulation (Model 1)**.

**Supplementary Table 8. Associations between physical frailty and the annual rate of system-specific chronic disease accumulation (Model 2)**.

**Supplementary Table 9. Associations between physical frailty and the annual rate of system-specific chronic disease accumulation (Model 3)**.

**Supplementary Table 10. Associations between physical frailty and the annual rate of chronic disease accumulation, stratified by gender**.

**Supplementary Table 11. Associations between physical frailty and the annual rate of chronic disease accumulation, stratified by age**.

**Supplementary Table 12. Sensitivity analysis of the associations between physical frailty and the annual rate of chronic disease accumulation, after excluding participants developing incident diseases during the first 3 years of follow-up**.

**Supplementary Table 13. Sensitivity analysis of the associations between physical frailty and the annual rate of chronic disease accumulation, restricted to participants with at least 5 years of follow-up**.

**Supplementary Figure 1. Association between physical frailty and the annual rate of total chronic disease accumulation according to baseline disease burden (Model 1)**. Linear mixed models were applied without adjustment for covariates (Model 1). Analysis was conducted across four groups: all participants, baseline disease-free, single baseline condition, and baseline multimorbidity. Data represent average predicted chronic disease counts with 95% confidence intervals.

**Supplementary Figure 2. Association between physical frailty and the annual rate of total chronic disease accumulation according to baseline disease burden (Model 2)**. Linear mixed models were adjusted for age, gender, race/ethnicity, education, and TDI (Model 2). Analysis was conducted across four groups: all participants, baseline disease-free, single baseline condition, and baseline multimorbidity. Data represent average predicted chronic disease counts with 95% confidence intervals. TDI: Townsend Deprivation Index.

